# Cumulative Prenatal Adversity and Fronto-Limbic White Matter Development Across Adolescence

**DOI:** 10.64898/2026.10.22.26366259

**Authors:** Lauren A. Costello, Hugh Garavan, Carolina Makowski, Jonathan Ahern, Alexandra S. Potter, Brie M. Reid, Micah E. Johnson, Matthew D. Albaugh, Priscila Dib Gonçalves

**Affiliations:** Department of Psychology, Northeastern University, Boston, MA; Institute for Cognitive and Brain Health, Northeastern University, Boston, MA; Department of Psychiatry, University of Vermont, Burlington, VT; Department of Family Medicine, David Geffen School of Medicine, University of California, Los Angeles, CA; Department of Psychiatry, University of California San Diego, San Diego, CA; Department of Cognitive Science, University of California San Diego, San Diego, CA; Department of Public Health and Health Sciences, Northeastern University, Boston, MA; Department of Psychiatry, Columbia University, New York, NY

## Abstract

**Importance:** The prenatal period is a critical window for intergenerational transmission of adversity, yet the neural substrates through which cumulative prenatal stress becomes biologically embedded in offspring brain development remain poorly understood. White matter microstructure is a candidate neurobiological substrate linking prenatal adversity to downstream emotional dysregulation.

**Objective:** To examine associations between cumulative adverse prenatal exposure (APE) burden and fronto-limbic white matter fractional anisotropy (FA) at baseline and across adolescent neurodevelopmental trajectories, and to test whether white matter FA mediates prenatal APE effects on emotional dysregulation.

**Design, Setting, and Participants:** Cohort study using data from the Adolescent Brain Cognitive Development (ABCD) Study™, Release 7.0, a diverse longitudinal cohort of US children recruited at ages 9 to 10 years across 21 sites. After diffusion tensor imaging quality control exclusions and retention of one twin or triplet per family, the final cross-sectional analytic sample comprised 10,483 participants. Longitudinal analyses included participants with data at one or more follow-up timepoints across four imaging waves.

**Exposures:** Cumulative APE burden, operationalized as a sum of six binary adverse prenatal exposures (unplanned pregnancy; early alcohol, tobacco, and cannabis use before pregnancy recognition; complicated pregnancy; complicated birth; range, 0-6) following Zhi et al. (2026).

**Main Outcomes and Measures:** Fractional anisotropy in 11 *a priori* fronto-limbic white matter tracts at baseline (ages approximately 10 years) and across four imaging timepoints (ages approximate 10, 12, 14, and 16 years). Exploratory outcomes included Child Behavior Checklist (CBCL) dysregulation profile and Emotion Regulation Questionnaire (ERQ) subscales.

**Results:** Higher prenatal APE burden was associated with lower baseline FA in left uncinate fasciculus (b = −0.00096; 95% CI, −0.00151 to −0.00042; FDR-corrected P = .006), corpus callosum (b = −0.00079; 95% CI, −0.00125 to −0.00032; FDR-corrected P = .006), and right uncinate fasciculus (b = −0.00071; 95% CI, −0.00120 to −0.00021; FDR-corrected P = .019). This main-effect association was stable across visits and robust to stringent adjustment for two complementary image-quality metrics (framewise displacement and frame censoring); prenatal burden was itself unassociated with image quality. Higher APE burden was additionally associated with steeper FA increases across adolescence, with a positive APE × Age interaction in 5 of 11 tracts (positive in direction across all 11); these trajectory effects were more sensitive to image quality, although the right uncinate fasciculus association survived stringent adjustment. Post hoc analyses indicated that pregnancy complications and prenatal cannabis exposure were the most consistent contributors. White matter FA partially mediated the association between prenatal APE burden and reactive emotional dysregulation, with no association with voluntary emotion regulation strategies.

**Conclusions and Relevance:** Cumulative prenatal APE burden was associated with lower fronto-limbic white matter FA —particularly in the uncinate fasciculus and corpus callosum—an enduring main-effect association that was stable across adolescence and robust to rigorous image-quality adjustment. Evidence for altered developmental trajectories was more sensitive to image quality and should be interpreted as a more tentative, complementary finding. These findings identify fronto-limbic white matter as a candidate neurobiological correlate of cumulative prenatal adversity, while underscoring that this pathway reflects a bounded set of prenatal exposures.

**Key Points:** *Question:* Is cumulative adverse prenatal exposure adversity associated with fronto-limbic white matter microstructure across adolescence?

*Findings:* In this cohort study of 10, 483 adolescents from the ABCD Study, higher adverse prenatal exposure burden was associated with lower fractional anisotropy in bilateral uncinate fasciculus and corpus callosum —a persistent main-effect association that was robust to rigorous image-quality assessment. Higher burden was additionally associated with steeper fractional anisotropy increases in 5 of 11 tracts, and white matter partially mediated associations with reactive emotional dysregulation.

*Meaning:* These findings identify fronto-limbic white matter microstructure as a candidate neurobiological correlate of cumulative prenatal adversity, consistent with early biological embedding through prenatal programming.

## Introduction

Adversity does not begin at birth. Maternal experiences during pregnancy, including stress, prenatal substance use, and obstetric complications, reach the developing fetus through shared gestational biology, shaping offspring neurodevelopment before the first postnatal breath^1,2^. This process, termed fetal programming or prenatal programming, links maternal adversity to lasting alterations in offspring stress physiology, immune function, and brain development through neuroendocrine, inflammatory, and epigenetic mechanisms^2–5^. Yet the neural substrates through which cumulative prenatal adverse exposures (APE) become biologically embedded in offspring brain development — and whether such embedding persists or evolves across adolescence — remain poorly characterized.

White matter microstructure is a compelling candidate substrate for these prenatal effects. Fractional anisotropy (FA), a diffusion tensor imaging index of white matter organization, increases across childhood and adolescence as experience-dependent myelination proceeds^6^. Fronto-limbic tracts, including the uncinate fasciculus, cingulate cingulum, corpus callosum, and superior longitudinal fasciculus, are central to emotion regulation and stress reactivity^7^. They mature protractedly, with the uncinate fasciculus among the last to reach full maturation in the third decade of life^8^. These tracts are also sensitive to early adversity: exposure is associated with fronto-limbic FA alterations in institutionalized^9^ and maltreated^10^ children, with timing of exposure shaping which tracts are most affected^11^. However, prior work has largely examined postnatal adversity or individual prenatal exposures in isolation^11–15^; whether cumulative prenatal adversity burden shapes fronto-limbic FA development across the full adolescent myelination window remains unknown^16^.

Large-scale longitudinal cohorts now make it possible to examine prenatal programming of brain development with substantial statistical power^17–19^. Zhi et al. (2026) demonstrated using data from the Adolescent Brain Cognitive Development (ABCD) Study that cumulative adverse prenatal exposures (APE) burden was associated with elevated and persistent psychopathology and accelerated cortical thinning across 36 of 68 cortical regions from ages 9 to 15^16^. Whether this same prenatal burden shapes white matter microstructural development, and whether effects reflect altered developmental trajectories rather than fixed baseline differences, has not been examined.

Here we characterize associations between cumulative APE burden and FA across 11 fronto-limbic tracts selected *a priori* using ABCD Study Data that follows youth up to age 16, incorporating two additional years beyond prior published ABCD analyses^16^. We hypothesized that higher prenatal APE burden would be associated with lower FA at baseline and with altered FA developmental trajectories across adolescence. As an exploratory analysis, we tested whether white matter FA mediated associations between prenatal APE burden and emotional dysregulation.

## Methods

### Sample

Data were drawn from the Adolescent Brain Cognitive Development (ABCD) Study™, Release 7.0, a longitudinal cohort study of children recruited at ages 9 to 10 years across 21 sites in the United States^20^. Because twins and higher-order multiples share a single intrauterine environment, their prenatal adverse exposure is largely identical — 91.6% of complete twin pairs had identical APE scores — making co-twins non-independent observations of the prenatal exposure. We therefore retained one randomly selected member per multiple-birth family. After DTI quality-control exclusions (retaining participants with zero failed DTI scans; N = 11,414), this yielded a cross-sectional analytic sample of 10 483 participants. Participants with missing twin-status data (n = 2509) were treated as singletons. Because non-twin siblings from the same families remained in the sample, all models included family (nested within site) as a random effect to account for residual familial clustering. For longitudinal analyses, participants with DTI data at 1 or more follow-up timepoints were included; DTI data were available at baseline (N = 10,960), 2-year follow-up (N = 7568), 4-year follow-up (N = 6016), and 6-year follow-up (N = 7183). Most participants (64%) contributed three or four timepoints (29,067 observations).

### Adverse Prenatal Exposures (APEs)

Cumulative adverse prenatal exposure (APE) burden was operationalized following Zhi et al. (2026)^16^, using 6 binary items from the ABCD prenatal history questionnaire: unplanned pregnancy, early alcohol, tobacco, and cannabis exposure (assessed as maternal use before knowledge of pregnancy), complicated pregnancy, and complicated birth. Items were summed to create a continuous APE burden score (range, 0-6; mean [SD], 1.50 [1.18]). The score was re-derived in Release 7.0, and the prevalence of each of the six exposures closely matched those reported by Zhi et al. (2026), confirming consistent operationalization (see Supplement).

### Neuroimaging

DTI data were acquired as part of the ABCD Study imaging acquisition protocol^21^ across 4 timepoints: baseline (mean [SD] age, 10.0 [0.6] years), 2-year follow-up (mean [SD] age, 12.1 [0.7] years), 4-year follow-up (mean [SD] age, 14.2 [0.7] years), and 6-year follow-up (mean age approximately 16 years). Diffusion-weighted images were processed using the ABCD Study’s standardized pipeline^22^, which includes eddy-current and motion correction, B0 distortion correction, and gradient nonlinearity correction. Fractional anisotropy (FA) was derived from the diffusion tensor, and tract-averaged FA values were extracted using AtlasTrack probabilistic atlas-based tractography^23^; tabulated tract-level FA from Release 7.0 was used for analysis. White matter tract visualizations were generated using the ggAtlasTrack R package^24^.

Tract-averaged FA was examined for 11 white matter tracts selected based on prior literature linking fronto-limbic white matter to stress exposure and emotion regulation^9,16,20^: bilateral uncinate fasciculus, bilateral cingulate cingulum, corpus callosum, bilateral parietal superior longitudinal fasciculus (pSLF), bilateral superior longitudinal fasciculus (SLF), and bilateral temporal superior longitudinal fasciculus (tSLF). Mean framewise displacement (MFD; excluding y-displacement) was computed across retained scans and used in sensitivity analyses. Frame censoring — the number of diffusion volumes removed during preprocessing for excessive motion or artifact, with higher counts indicating poorer quality — was derived from the ABCD DTI quality-control pipeline and used as a time-varying image-quality index in sensitivity analyses. MRI scanner software version (12 levels combining device model and software release) was included as a fixed-effect covariate in all models.

### Dysregulation Profile

The Child Behavior Checklist^25,26^ (CBCL) dysregulation profile was computed as the sum of raw scores from 3 syndrome subscales: Attention Problems, Aggressive Behavior, and Anxious/Depressed, indexing what has been conceptualized as involuntary reactive emotional dysregulation.

### Emotion Regulation Questionnaire (ERQ)

ERQ^27^ cognitive reappraisal and expressive suppression subscales, available at the 2-year follow-up, were examined as an exploratory specificity analysis.

### Statistical Analyses

All analyses were conducted in R version 4.5.1^28^ using linear mixed-effects regression models^29,30^, with Satterthwaite approximation for degrees of freedom. Fixed effects covariates included child age, sex, race, ethnicity, caregiver educational attainment, and MRI scanner software version. Random effects accounted for the nested data structure: participants within families within sites.

#### Cross-sectional primary analyses

For each of the 11 tracts of interest, we regressed baseline FA on APE burden and covariates. FDR correction was applied across tracts (Benjamini-Hochberg)^31^.

#### Longitudinal primary analyses

Trajectories were modeled with linear mixed-effects models using the lme4 package^29^, with Satterthwaite degrees of freedom via lmerTest^30^. Mixed-effects models use all available observations under a missing-at-random assumption; participants with a single timepoint therefore contribute to estimation of the intercept, while those with repeated measures inform the age slope. FA was z-scored within tract and mean-centered age included as a random slope, and the APE × Age interaction was the primary test of trajectory moderation. To address known nonlinearity in FA maturation, we additionally tested whether a quadratic age term (age²) and its interaction with APE burden (APE × age²) improved model fit; the APE × age² term assessed whether burden moderated nonlinear trajectory components. Likelihood-ratio tests confirmed that including a random age slope significantly improved model fit over a random-intercept-only specification in all 11 tracts (all P < 10⁻²²), with lower AIC and BIC for the random-slope model in every tract; random slopes were therefore retained.

#### Exploratory analyses

Mediation analyses tested whether tract FA mediated APE-dysregulation associations using bootstrap resampling (1000 iterations)^32^. ERQ subscales were regressed on APE using the same covariate structure.

#### Sensitivity analyses

All cross-sectional models were re-run with MFD as an additional covariate. Supplementary analyses examined polygenic risk for total problems on tract FA in participants of European ancestry (n ≈ 6270), with APE and the top 10 genetic principal components as covariates to assess whether genetic risk related to FA independently of prenatal exposure.

## Results

### Sample Characteristics

The final cross-sectional analytic sample comprised 10 483 participants at the baseline visit (mean [SD] age, 9.94 [0.63] years; 52.3% female; Table 1). The APE burden score ranged from 0 to 6 (mean [SD], 1.50 [1.18]).

**Table 1.** Baseline sample characteristics stratified by adverse prenatal exposure (APE)

| Characteristic | **Total**<br>**(n = 10 483)** | **APE = 0**<br>**(n = 2475)** | **APE = 1**<br>**(n = 3462)** | **APE = 2**<br>**(n = 2634)** | **APE = 3+**<br>**(n = 1912)** | **Statistic** | ***p*** |
|---|---|---|---|---|---|---|---|
| Female, N (%) | 5480 (52.3%) | 1258 (50.8%) | 1805 (52.1%) | 1418 (53.8%) | 999 (52.5%) | χ<sup>2</sup> = 4.61 | .203 |
| Age, mean (SD), y | 9.94 (0.63) | 9.92 (0.63) | 9.93 (0.63) | 9.95 (0.63) | 9.97 (0.63) | F = 2.70 | .044 |
| Race, N (%) |  |  |  |  |  |  |  |
| White | 6904 (65.9%) | 1540 (62.2%) | 2361 (68.1%) | 1794 (68.0%) | 1209 (63.6%) | χ<sup>2</sup> = 116.74 | < .001 |
| Black or African American | 1689 (16.1%) | 414 (16.7%) | 527 (15.2%) | 398 (15.1%) | 350 (18.4%) |  |  |
| Asian | 238 (2.3%) | 103 (4.2%) | 66 (1.9%) | 50 (1.9%) | 19 (1.0%) |  |  |
| Other | 57 (0.5%) | <20<sup>a</sup> | <20<sup>a</sup> | <20<sup>a</sup> | <20<sup>a</sup> |  |  |
| American Indian / Alaskan Native | 15 (0.1%) | <10<sup>a</sup> | <10<sup>a</sup> | <10<sup>a</sup> | <10<sup>a</sup> |  |  |
| More than One Race | 1147 (10.9%) | 266 (10.7%) | 347 (10.0%) | 286 (10.8%) | 243 (13.0%) |  |  |
| Unknown or not reported | 432 (4.1%) | 133 (5.4%) | 146 (4.2%) | 97 (3.7%) | 56 (2.9%) |  |  |
| Ethnicity, N (%) |  |  |  |  |  |  |  |
| Hispanic/Latino | 2336 (21.4%) | 530 (21.5%) | 790 (22.9%) | 567 (21.6%) | 349 (18.4%) | χ<sup>2</sup> = 15.01 | .002 |
| Non-Hispanic/Latino | 8206 (78.6%) | 1939 (78.5%) | 2657 (77.1%) | 2061 (78.4%) | 1549 (81.6%) |  |  |
| Caregiver educational attainment, N (%) |  |  |  |  |  |  |  |
| Up to high school (no diploma) | 544 (5.2%) | 135 (5.5%) | 208 (6.0%) | 136 (5.2%) | 65 (3.4%) | χ<sup>2</sup> = 167.50 | < .001 |
| High school/GED | 1012 (9.7%) | 255 (10.3%) | 320 (9.2%) | 236 (9.0%) | 201 (10.6%) |  |  |
| Some college | 2713 (25.9%) | 513 (20.8%) | 829 (23.9%) | 695 (26.4%) | 676 (35.6%) |  |  |
| Bachelor’s degree | 2621 (25.0%) | 623 (25.2%) | 877 (25.3%) | 668 (25.4%) | 453 (23.8%) |  |  |
| Graduate school or professional degree | 3579 (34.2%) | 945 (38.2%) | 1231 (35.5%) | 898 (34.1%) | 505 (26.6%) |  |  |
| Scanner, N (%) |  |  |  |  |  |  |  |
| GE | 2801 (26.7%) | 711 (28.7%) | 904 (26.1%) | 702 (26.6%) | 484 (25.3%) | χ<sup>2</sup> = 12.48 | .052 |
| Philips | 1269 (12.1%) | 288 (11.6%) | 456 (13.2%) | 308 (11.7%) | 217 (11.3%) |  |  |
| Siemens | 6413 (61.2%) | 1478 (59.7%) | 2106 (60.8%) | 1628 (61.8%) | 1201 (62.8%) |  |  |
| White Matter FA, mean (SD) |  |  |  |  |  |  |  |
| Uncinate LH | 0.44 (0.04) | 0.45 (0.04) | 0.44 (0.04) | 0.44 (0.04) | 0.44 (0.05) | F = 3.59 | .013 |
| Uncinate RH | 0.42 (0.04) | 0.42 (0.04) | 0.42 (0.04) | 0.42 (0.04) | 0.42 (0.04) | F = 3.11 | .025 |
| CgC LH | 0.49 (0.06) | 0.49 (0.05) | 0.49 (0.06) | 0.49 (0.06) | 0.49 (0.06) | F = 4.25 | 0.005 |
| CgC RH | 0.44 (0.05) | 0.44 (0.05) | 0.44 (0.05) | 0.44 (0.05) | 0.44 (0.06) | F = 1.07 | .362 |
| Corpus callosum | 0.57 (0.04) | 0.57 (0.04) | 0.57 (0.04) | 0.57 (0.04) | 0.57 (0.04) | F = 3.35 | .018 |
| pSLF LH | 0.47 (0.04) | 0.47 (0.03) | 0.46 (0.04) | 0.47 (0.03) | 0.46 (0.04) | F = 1.91 | .125 |
| pSLF RH | 0.47 (0.04) | 0.47 (0.03) | 0.47 (0.04) | 0.47 (0.04) | 0.47 (0.04) | F = 1.17 | .319 |
| SLF LH | 0.49 (0.04) | 0.49 (0.03) | 0.49 (0.04) | 0.49 (0.03) | 0.49 (0.04) | F =1.17 | .320 |
| SLF RH | 0.48 (0.04) | 0.48 (0.03) | 0.48 (0.04) | 0.48 (0.03) | 0.48 (0.04) | F = 1.04 | .372 |
| tSLF LH | 0.51 (0.04) | 0.51 (0.03) | 0.50 (0.04) | 0.51 (0.03) | 0.50 (0.04) | F = 0.83 | .476 |
| tSLF RH | 0.49 (0.04) | 0.49 (0.03) | 0.49 (0.04) | 0.49 (0.03) | 0.49 (0.04) | F = 0.66 | .578 |
| Prenatal exposures, N (%) |  |  |  |  |  |  |  |
| Unplanned pregnancy | 3620 (35.0%) | 0 (0.0%) | 954 (27.6%) | 1364 (52.0%) | 1302 (68.8%) | NA | NA |
| Alcohol exposure | 2568 (26.0%) | 0 (0.0%) | 590 (18.1%) | 845 (33.4%) | 1133 (61.0%) | NA | NA |
| Marijuana exposure | 620 (5.8%) | 0 (0.0%) | 30 (0.9%) | 151 (5.8%) | 439 (23.6%) | NA | NA |
| Nicotine exposure | 1417 (13.9%) | 0 (0.0%) | 170 (5.0%) | 399 (15.3%) | 848 (44.9%) | NA | NA |
| Birth complication | 2672 (25.5%) | 0 (0.0%) | 571 (16.5%) | 919 (34.8%) | 1182 (62.1%) | NA | NA |
| Pregnancy complication | 4240 (40.4%) | 0 (0.0%) | 1151 (33.2%) | 1598 (60.6%) | 1491 (78.4%) | NA | NA |

**Note.** *Values are mean (SD) or N (%). APE = adverse prenatal exposure count (0–6); participants are grouped by number of prenatal exposures. χ<sup>2</sup> tests used for categorical variables; one-way ANOVA for continuous variables. CgC = cingulate cingulum; FA = fractional anisotropy; LH = left hemisphere; RH = right hemisphere; pSLF = parietal superior longitudinal fasciculus; SLF = superior longitudinal fasciculus; tSLF = temporal superior longitudinal fasciculus. Prenatal exposure cells show 0 (0.0%) for APE = 0 by definition, as no exposures were endorsed in that group.* <sup>*a*</sup> *Per Adolescent Brain Cognitive Development (ABCD) Study data-use requirements, cell frequencies of 1 to 9 are reported as “<10” to protect participant privacy; a second cell in each affected column is reported as “<20” to prevent recovery of the suppressed value by subtraction. χ<sup>2</sup> statistics were computed on the complete (unsuppressed) data.*

### APE and Baseline White Matter FA

Higher APE burden was associated with lower baseline FA across all 11 tracts. After FDR correction, effects were significant in the uncinate fasciculus bilaterally and corpus callosum (all P_FDR ≤ .019); the left cingulate cingulum and right pSLF were nominally significant but did not survive correction (Figure 1; Table 2).

**Figure 1.**
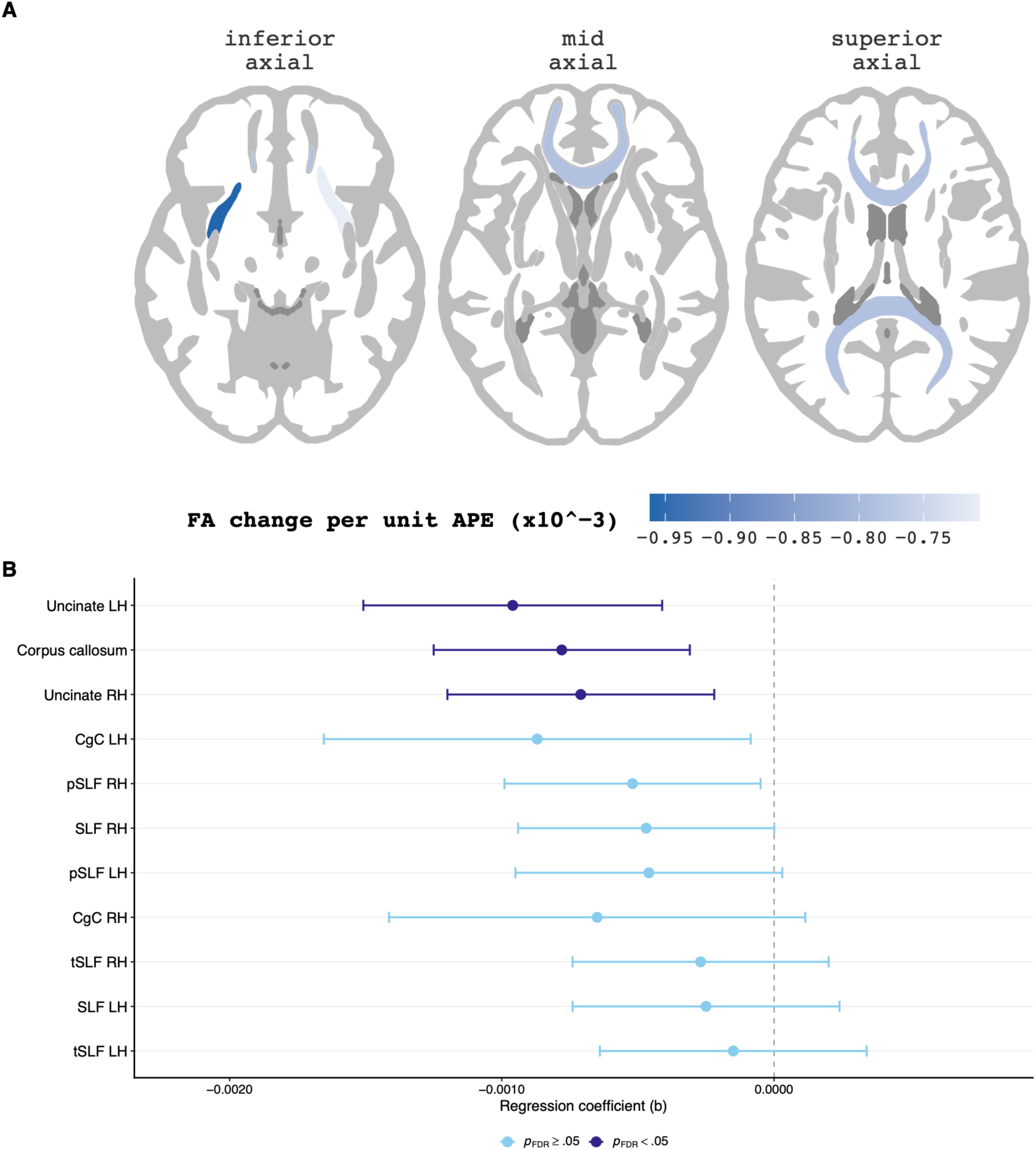
Associations Between Cumulative Adverse Prenatal Exposure Burden and Baseline White Matter Fractional Anisotropy. A, Axial views (inferior, mid, and superior) displaying the association between cumulative adverse prenatal exposure (APE) burden and baseline fractional anisotropy (FA) across the 11 a priori white matter tracts; darker shading indicates more negative associations (FA change per unit APE burden). B, Forest plot of unstandardized regression coefficients (b) and 95% CIs for the association between cumulative APE burden and baseline FA (N = 10 483). Models included age, sex, race, ethnicity, caregiver educational attainment, and MRI scanner software version as covariates, with random effects for site and family. Tracts are ordered by effect size. Dark purple indicates FDR-corrected P < .05; light blue indicates FDR-corrected P ≥ .05. CgC indicates cingulate cingulum; FA, fractional anisotropy; FDR, false discovery rate; LH, left hemisphere; pSLF, parietal superior longitudinal fasciculus; RH, right hemisphere; SLF, superior longitudinal fasciculus; tSLF, temporal superior longitudinal fasciculus.

**Table 2.** Associations between APE and white matter FA across 11 a priori tracts.

| **Tract** | ***B (95% CI)*** | ***P*** | ***P*<sup>FDR</sup>** |
|---|---|---|---|
| *Uncinate fasciculus (LH)* | −0.00096 (−0.00151 to −0.00042) | .001 | .006** |
| *Corpus callosum* | −0.00079 (−0.00125 to −0.00032) | .001 | .006** |
| *Uncinate fasciculus (RH)* | −0.00071 (−0.00120 to −0.00021) | .005 | .019* |
| *CgC (LH)* | −0.00087 (−0.00166 to −0.00009) | .030 | .068 |
| *pSLF (RH)* | −0.00053 (−0.00100 to −0.00005) | .029 | .068 |
| *SLF (RH)* | −0.00048 (−0.00095 to −0.00001) | .047 | .085 |
| *pSLF (LH)* | −0.00046 (−0.00095 to 0.00002) | .065 | .102 |
| *CgC (RH)* | −0.00066 (−0.00143 to 0.00010) | .096 | .131 |
| *SLF (LH)* | −0.00026 (−0.00074 to 0.00022) | .302 | .332 |
| *tSLF (RH)* | −0.00028 (−0.00075 to 0.00019) | .237 | .305 |
| *tSLF (LH)* | −0.00016 (−0.00065 to 0.00034) | .547 | .547 |

\*P<sup>FDR</sup> < .05. \*\*P<sup>FDR</sup> < .01. Models estimated using linear mixed-effects regression with Satterthwaite degrees of freedom. Fixed effects covariates: age, sex, race, ethnicity, caregiver education, MRI scanner software version (12 levels). Random effects: (1 \| site / family\_id). b indicates unstandardized regression coefficient; CgC = cingulate cingulum; CI, confidence interval; APE, adverse prenatal exposure; FA, fractional anisotropy; FDR, false discovery rate; LH, left hemisphere; PSLF, parietal superior longitudinal fasciculus; RH, right hemisphere; SLF, superior longitudinal fasciculus; tSLF, temporal superior longitudinal fasciculus.

### APE and White Matter Developmental Trajectories

FA increased with age across all 11 tracts (all P < .0001), consistent with active myelination. Across all available visits, higher APE burden was associated with lower FA generally, with significant main effects in the corpus callosum (P = .004) and left uncinate fasciculus (P = .011). This main-effect association between prenatal burden and white matter was stable across visits. A significant positive APE × Age interaction was additionally observed in 5 of 11 tracts after FDR correction (bilateral uncinate, left cingulate cingulum, right pSLF, right SLF; Figure 2; Table 3). Critically, the interaction was positive in the same direction across all 11 tracts, indicating that higher APE burden was consistently associated with a steeper rate of FA increase across adolescence (Supplementary Figure 1).

**Figure 2.**
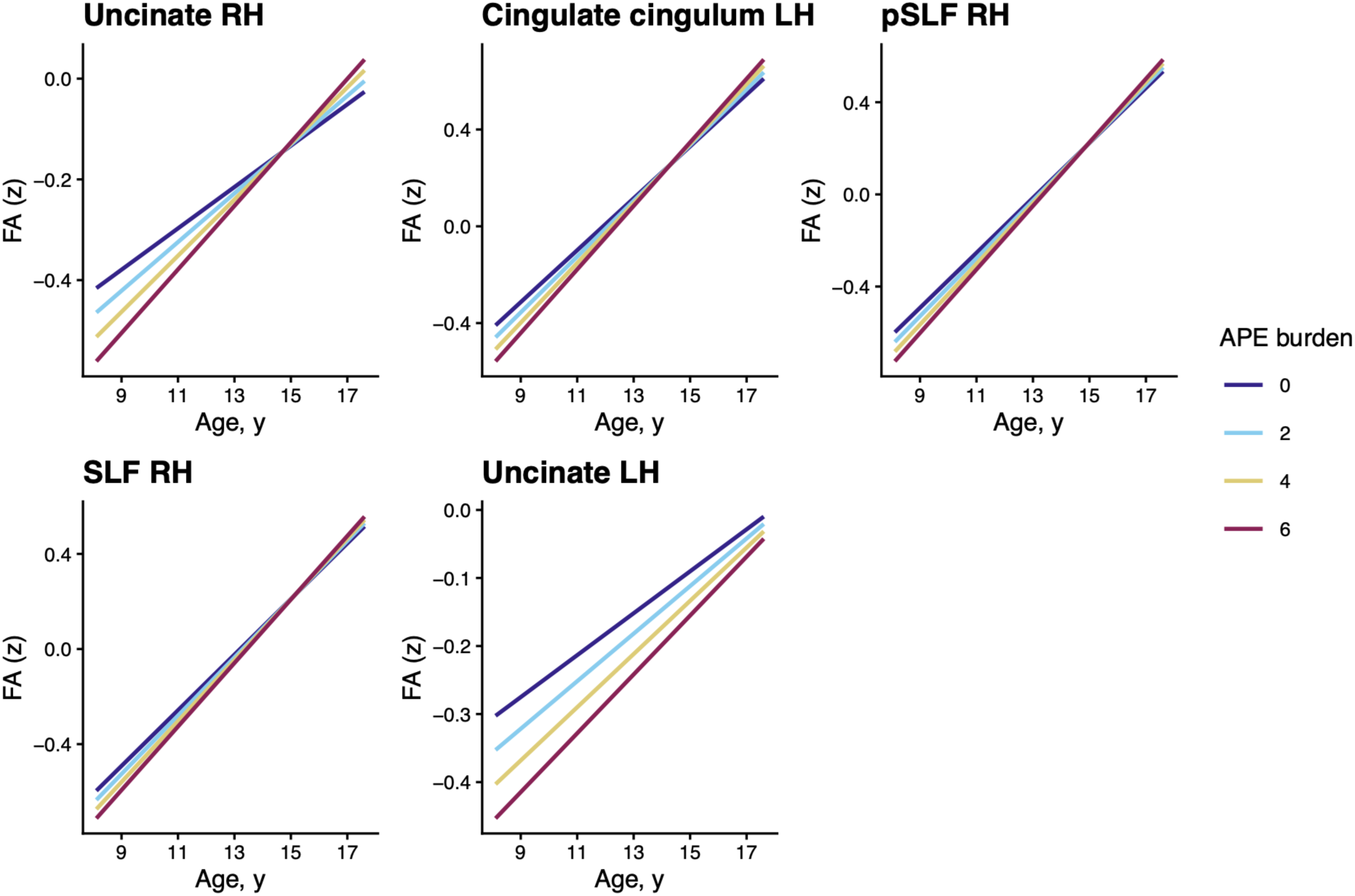
Prenatal Adverse Prenatal Exposure Burden and White Matter Fractional Anisotropy Developmental Trajectories Across Adolescence. Model-predicted FA trajectories across adolescence (ages ∼9–17 years) for the five fronto-limbic tracts with significant APE × age interactions. Predictions are derived from linear mixed-effects models treating cumulative APE burden as a continuous predictor; FA was z-scored within tract. Trajectories are displayed at representative burden levels (0, 2, 4, and 6 adverse prenatal exposures), with models estimated across the full burden range (0–6). For tracts in which a quadratic age term significantly improved model fit (uncinate fasciculus RH, pSLF RH, SLF RH), quadratic trajectories are shown; for the remaining tracts (cingulate cingulum LH, uncinate fasciculus LH), first-order linear trajectories are shown. CgC indicates cingulate cingulum; FA, fractional anisotropy; pSLF, parietal superior longitudinal fasciculus; RH, right hemisphere; LH, left hemisphere; SLF, superior longitudinal fasciculus.

**Table 3.** APE burden and white matter FA developmental trajectories (APE × Age interaction)

| **Tract** | ***B (95% CI)*** | ***P*** | ***P*<sup>FDR</sup>** |
|---|---|---|---|
| *Uncinate fasciculus (RH)* | 0.0040 (0.0018 to 0.0062) | .0005 | .005 ** |
| *CgC (LH)* | 0.0040 (0.0014 to 0.0065) | .003 | .014 * |
| *pSLF (RH)* | 0.0030 (0.0007 to 0.0053) | .009 | .035 * |
| *SLF (RH)* | 0.0028 (0.0004 to 0.0051) | .022 | .049 * |
| *Uncinate fasciculus (LH)* | 0.0026 (0.0004 to 0.0048) | .022 | .049 * |
| *pSLF (LH)* | 0.0024 (0.0000 to 0.0049) | .049 | .090 |
| *Corpus callosum* | 0.0021 (−0.0001 to 0.0043) | .063 | .095 |
| *SLF (LH)* | 0.0022 (−0.0002 to 0.0046) | .069 | .095 |
| *tSLF (LH)* | 0.0020 (−0.0004 to 0.0043) | .100 | .122 |
| *tSLF (RH)* | 0.0017 (−0.0005 to 0.0038) | .133 | .146 |
| *CgC (RH)* | 0.0016 (−0.0007 to 0.0039) | .169 | .169 |

\*P<sup>FDR</sup> < .05. \*\*P<sup>FDR</sup> < .01. All APE × Age interactions were positive in direction. Linear mixed-effects (growth curve) models with FA z-scored within tract; coefficients represent the change in annual FA slope (SD units) per additional prenatal exposure. Fixed effects: APE burden × mean-centered age, plus sex, caregiver educational attainment, race, ethnicity, and MRI scanner software version (12 levels). Random effects: random intercept and slope for age at the participant level, with participants nested within families and study sites [(1 + age \| participant) + (1 \| site/family)]. Four imaging timepoints (ages ≈10-16 years). APE indicates adverse prenatal exposure; b, unstandardized coefficient; CgC = cingulate cingulum; CI, confidence interval; FA, fractional anisotropy; FDR, false discovery rate; LH, left hemisphere; pSLF, parietal superior longitudinal fasciculus; RH, right hemisphere; SLF, superior longitudinal fasciculus; tSLF, temporal superior longitudinal fasciculus.

Consistent with known nonlinearity in FA maturation, a quadratic age term significantly improved fit in four tracts (corpus callosum, right pSLF, right SLF, right TSLF; all P < .01, with concordant reductions in AIC and BIC). However, the APE × age² interaction was nonsignificant in all 11 tracts (all P ≥ .32), indicating that prenatal burden did not moderate nonlinear trajectory components; the linear APE × Age interaction remained significant in the primary tracts after inclusion of quadratic terms. The burden-related trajectory effect was therefore best characterized as linear.

### Sensitivity and Exploratory Analyses

#### Image-quality sensitivity analyses

To assess whether associations reflected diffusion image quality rather than prenatal adversity, we conducted complementary quality-control analyses using two metrics: mean framewise displacement (MFD) and frame censoring. Prenatal APE burden was not associated with the number of censored frames at baseline (b = 0.03; P = .31), indicating that children with higher adversity burden did not have poorer baseline data quality. Baseline associations were robust to adjustment for both metrics: the three FDR-significant associations (bilateral uncinate fasciculus and corpus callosum) were essentially unchanged with MFD included (Supplementary Table 1) and remained significant after adjusting for frame censoring (Supplementary Table 2). In repeated-measures models, the APE × Age interaction in the right uncinate fasciculus survived adjustment for time-varying frame censoring (P_FDR = .04), though the remaining four interactions attenuated to non-significance. The APE burden main effect remained significant under censoring adjustment (corpus callosum, left uncinate fasciculus; Supplementary Table 2), confirming that this stable association is robust to image quality.

### Maternal depression

Associations were robust to adjustment for maternal depression history: bilateral uncinate and corpus callosum effects were essentially unchanged (<6% attenuation of effect sizes) and remained FDR-significant, and trajectory interactions were likewise unchanged (<0.3% shift), with consistent results across full-sample and complete-case analyses (Supplementary Table 3).

### Mediation / specificity

Bootstrapped mediation analyses identified significant indirect effects of prenatal APE burden on CBCL dysregulation through FA in the bilateral uncinate fasciculus and corpus callosum, with significant residual direct effects, indicating partial mediation (Supplementary Table 6 and Figure 2). In a post-hoc analysis, prenatal APE burden was not associated with cognitive reappraisal (P = .63) or expressive suppression (P = .39), suggesting specificity to reactive dysregulation. Polygenic risk for total problems was nominally associated with lower left cingulate cingulum FA, independent of APE burden (Supplementary Table 4).

### Exposure-specific (domain) analyses

Testing each of the six APE domains individually, pregnancy complications were the most consistent contributor to lower baseline FA (significant across the corpus callosum and bilateral uncinate fasciculus), whereas prenatal cannabis exposure showed the largest per-exposure effects; for trajectories, cannabis showed the most consistent positive APE × Age interactions across the five primary tracts. Given the low prevalence of cannabis exposure (n = 620; ≈6%), these findings are exploratory and hypothesis-generating (Supplementary Table 5).

## Discussion

In the largest longitudinal study of prenatal adversity and white matter microstructure to date, we show that cumulative prenatal adversity is associated with lower fronto-limbic white matter FA in the bilateral uncinate fasciculus and corpus callosum — a developmentally stable association robust to rigorous image-quality adjustment. Higher burden was additionally associated with steeper FA increases across ages 10 to 16, interpreted as a secondary, more tentative finding, though the right uncinate fasciculus interaction survived stringent adjustment. These findings extend those of Zhi et al. (2026), which demonstrated APE burden effects on cortical thinning trajectories, to white matter microstructure, consistent with evidence from other large-scale developmental cohorts^14,33^.

### Intergenerational transmission via prenatal programming

A central contribution of the present study is situating prenatal adversity within an intergenerational transmission framework, in which maternal experiences reach the developing child’s brain before birth and leave an enduring neurobiological signature. Prenatal adversity associations with lower FA that persists across development, rather than emerging or resolving within this window, is consistent with early biological embedding. Our exposure captures a bounded set of prenatal experiences rather than the full range of maternal adversity, and we did not assess the biological mediators of transmission; thus, these findings document an association consistent with intergenerational programming rather than its mechanism. The partial mediation of prenatal APE effects through white matter FA converges with evidence that global white matter microstructure indexes general psychopathology independent of specific symptom dimensions^34^, positioning fronto-limbic FA as a candidate transdiagnostic substrate of risk. Future work integrating biological mediators, genetic risk, and prenatal programming may clarify these mechanisms^35^.

### Cross-sectional findings

The cross-sectional associations replicate and extend prior findings in smaller samples^12,36,37^. The uncinate fasciculus is particularly implicated given its role in threat appraisal and stress regulation; reduced uncinate FA follows prenatal maternal stress and postnatal adversity and tracks emotional dysregulation and internalizing risk^13–15,38,39^. The association between higher APE burden and lower corpus callosum FA aligns with ABCD evidence of prenatal effects on brain structure^16,19^, extending this literature from cortical morphology to white matter microstructure. These effects appear well before adolescence: prenatal social disadvantage is linked to reduced neonatal white matter^40^, and prenatal stress to altered white matter from infancy onward^14^, suggesting prenatal alterations that are subsequently remodeled across development.

### Developmental trajectories

Beyond the stable baseline association, higher APE burden was associated with steeper FA increases across adolescence, converging with emerging cross-cohort evidence. In the Generation R study, prenatal depression exposure was associated with lower childhood FA followed by faster increases across ages 7 to 15 — a “catch-up” pattern in overlapping tracts^33^ — which our finding mirrors. That this lower-baseline, steeper-increase signature now appears across two large, independent cohorts with different exposures and non-overlapping tract-identification pipelines is encouraging; however, under adjustment for time-varying image quality the interaction attenuated to a single FDR-significant tract. Notably, the right uncinate fasciculus interaction survived this adjustment — a tract whose orbitofrontal connectivity is consistent with its role in emotional and behavioral dysregulation^41^. The burden main effect, by contrast, remained significant regardless of quality adjustment, indicating that the stable association, not its change across age, is the more robust finding.

### Mediation and behavioral relevance

Partial mediation through uncinate and callosal FA positions fronto-limbic white matter as one of multiple pathways in prenatal adversity-related behavioral risk. APE burden was associated with reactive emotional dysregulation (CBCL) but not voluntary ERQ strategies, a specificity consistent with prenatal programming of subcortical-limbic circuits supporting automatic stress reactivity^1^, whereas voluntary ERQ strategies draw upon separately-developing prefrontal systems^42^.

### Strengths, limitations, and future directions

Strengths include the largest longitudinal sample to date (N = 10 483), four imaging timepoints spanning ages 10 to 16, rigorous covariate control, the validated APE burden score enabling direct comparison with Zhi et al. (2026)^16^. The ABCD Study further provides a sociodemographically diverse cohort with the longest span of repeated pediatric neuroimaging available. Modeling family-level clustering via mixed effects let us retain siblings rather than excluding them, preserving statistical power while handling family-based dependence^43^.

Limitations include retrospective caregiver report of prenatal exposures, which may introduce recall bias (particularly where the reporting caregiver was not the biological mother, although such misreporting would most likely bias associations toward the null); the inability to compare effects across gestational timing windows, as substance exposures were assessed only before pregnancy recognition; and the non-specificity of FA as an index of microstructural brain properties. More specific markers such as myelin water fraction or magnetization transfer imaging^6^ would provide additional interpretation of cellular properties. Relatedly, tract-averaged FA collapses substantial within-tract spatial variation, and localized effects may be obscured by whole-tract summaries. We also examined a priori fronto-limbic tracts without non-fronto-limbic control tracts or whole-brain analyses; the anatomical specificity of these associations therefore warrants further examination. The cross-sectional mediation design precludes causal inference. Although the right uncinate fasciculus APE × Age interaction survived stringent adjustment for time-varying image quality, the remaining trajectory associations attenuated, and we therefore interpret the age-related trajectory effects as more tentative than the stable baseline association. We adjusted for caregiver educational attainment but not household income, leaving residual socioeconomic confounding possible. Genetic confounding is a further limitation; future work integrating polygenic scores with prenatal programming models may disentangle heritable from environmentally mediated pathways^35,44^. We focused on maternal prenatal exposures, consistent with our gestational-biology framework; paternal contributions to offspring white matter were not examined and warrant future study.

Finally, approximately a decade elapsed between the prenatal exposures assessed here and the first neuroimaging assessment. Because prenatal adversity frequently co-occurs with, and predicts, postnatal adversity, we cannot fully disentangle prenatal programming from the contribution of correlated postnatal experiences, which we did not measure. However, several features of the design support a prenatal contribution: the exposures are temporally prior to any postnatal influence; the near-identical APE scores within twin pairs indicate they operate as shared gestational exposures; and, using this same exposure measure in ABCD, Zhi et al. demonstrated that APE associations with psychopathology and cortical thinning persisted in within-family sibling comparisons discordant for APE burden^16^, attenuating shared familial and postnatal confounding. Definitively isolating the prenatal window nonetheless requires longitudinal cohorts with neuroimaging from infancy, such as the HEALthy Brain and Child Development (HBCD) Study, which can measure brain structure before postnatal adversity accumulates.

## Conclusions

Cumulative prenatal APE burden was associated with lower fronto-limbic FA at the start of adolescence, an association whose main effect was developmentally stable and robust to adjustment for image quality. These findings identify fronto-limbic white matter microstructure, particularly the uncinate fasciculus and corpus callosum, as a candidate neurobiological correlate of cumulative prenatal adversity and a candidate pathway in its intergenerational transmission, while underscoring that this pathway reflects a bounded set of prenatal exposures. Prenatal screening and early intervention may represent important targets for supporting healthy neurodevelopment following prenatal adversity.

## Data Availability

Data used in this study are from the Adolescent Brain Cognitive Development (ABCD) Study and are available through the NIMH Data Archive (NDA). Access requires a data use agreement; qualified researchers can apply at https://nda.nih.gov/abcd. The authors are not permitted to share the data directly.

## ADDITIONAL CONTRIBUTIONS

Data used in the preparation of this article were obtained from the Adolescent Brain Cognitive Development™ (ABCD) Study, held in the NIH Brain Development Cohorts Data Sharing Platform. This is a multisite, longitudinal study designed to recruit more than 10,000 children aged 9–10 and follow them over 10 years into early adulthood. The ABCD Study® is supported by the National Institutes of Health and additional federal partners under award numbers U01DA041048, U01DA050989, U01DA051016, U01DA041022, U01DA051018, U01DA051037, U01DA050987, U01DA041174, U01DA041106, U01DA041117, U01DA041028, U01DA041134, U01DA050988, U01DA051039, U01DA041156, U01DA041025, U01DA041120, U01DA051038, U01DA041148, U01DA041093, U01DA041089, U24DA041123, and U24DA041147. A full list of supporters is available at https://abcdstudy.org/federal-partners.html. ABCD Consortium investigators designed and implemented the study and/or provided data but did not necessarily participate in the analysis or writing of this report. This manuscript reflects the views of the authors and may not reflect the opinions or views of the NIH or ABCD Consortium investigators. We also extend our thanks to the participants and families involved with the ABCD Study, and to the Scientific Training in Addiction Research Techniques (START) Program — in particular Skye Bristol, Nicholas Allgaier, and Sarahjane Dube — which made this paper possible.

## DATA SHARING STATEMENT

The ABCD data repository grows and changes over time. The ABCD data used in this report came from NBDC Digital Object Identifier https://doi.org/10.82525/8f3w-5260. DOIs can be found at https://www.nbdc-datahub.org/abcd-study.

