## Supplemental Figures and Tables for "Cumulative Prenatal Adversity and Fronto-Limbic White Matter Development Across Adolescence"

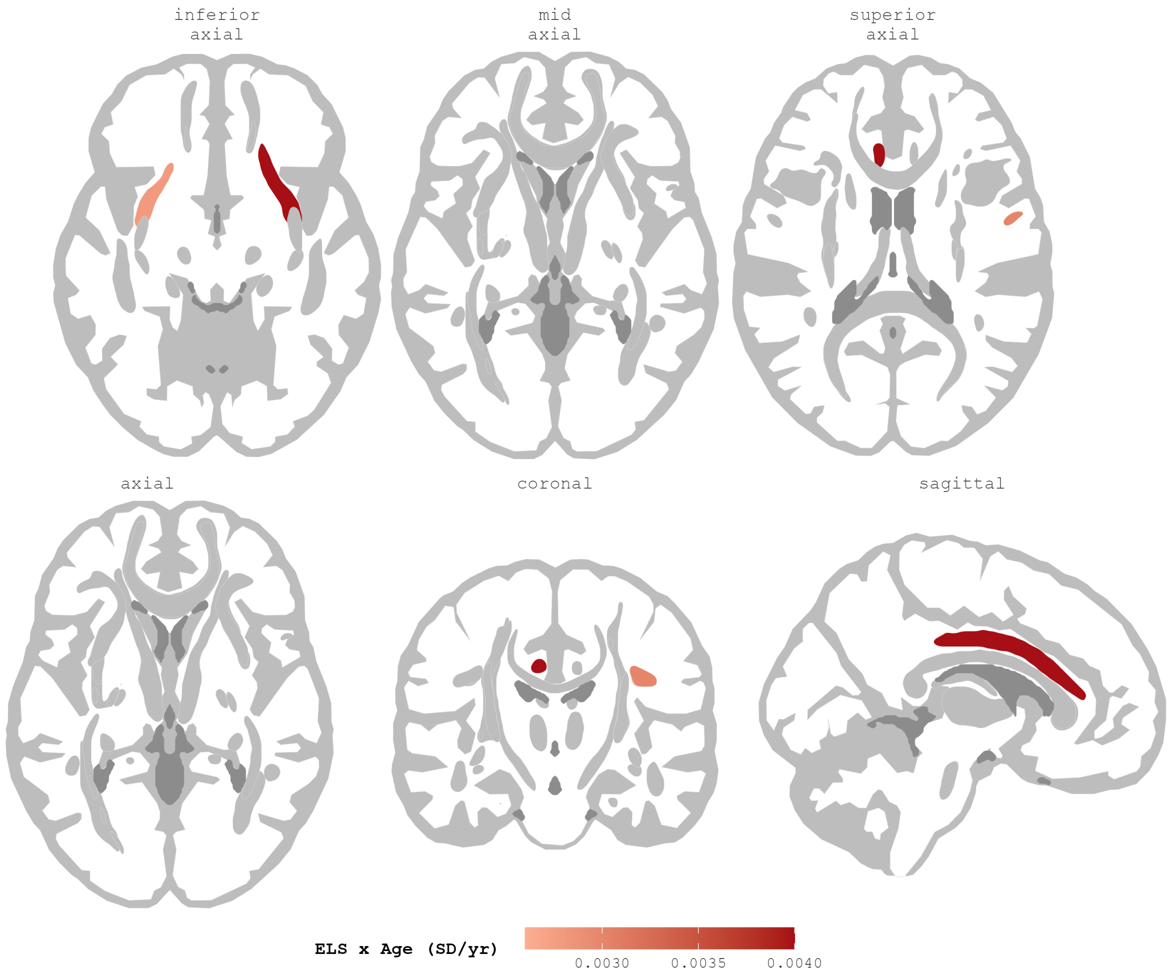


**Supplementary Figure 1. Cumulative Adverse Prenatal Exposure and Fronto-Limbic White Matter Developmental Trajectories.**

Tract-level APE burden × age interactions from linear mixed-effects growth-curve models across 4 imaging timepoints (ages ≈10-16 years), displayed on the AtlasTrack atlas in 2 complementary view sets: 3 axial planes (top row) and an orthogonal triptych (axial, coronal, sagittal; bottom row), so that each significant tract is legible in at least 1 plane. Color indicates the standardized APE × age interaction coefficient (change in annual FA slope, in SD units, per additional prenatal exposure); the interaction was positive (red) in all significant tracts, indicating steeper FA increases across adolescence with higher APE burden. Five tracts survived FDR correction:

right uncinate fasciculus (b = 0.0040; *P*^FDR^ = .005), left cingulate (b = 0.0040; *P*^FDR^ = .014), right parietal superior longitudinal fasciculus (b = 0.0030; *P*^FDR^ = .035), left uncinate fasciculus (b = 0.0028; *P*^FDR^ = .049), and right superior longitudinal fasciculus (b = 0.0026; *P*^FDR^ = .049). Tracts not surviving FDR correction are shown in gray. Models were adjusted for child age, sex, race, ethnicity, caregiver educational attainment, and MRI scanner software version, with random effects for participant-level age slopes and participants nested within families within sites.

c

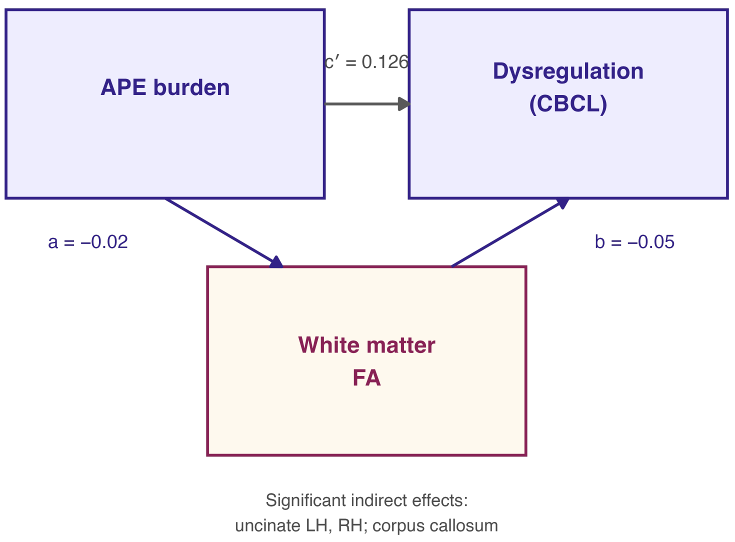


**Supplementary Figure 2. White Matter Fractional Anisotropy as a Mediator of the Association Between APE Burden and Emotional Dysregulation.**

Path diagram of bootstrapped mediation models (1 000 iterations) with APE burden as the predictor, white matter FA as the mediator, and CBCL dysregulation profile as the outcome. Path a = APE → FA; path b = FA → dysregulation (controlling for APE); path c′ = direct effect. Representative coefficients are averaged across the three tracts with significant indirect effects (left and right uncinate fasciculus and corpus callosum). Total effect of APE on dysregulation: b = 0.131 (SE, 0.008; $P < 10^{-54}$). APE indicates adverse prenatal exposure; CBCL indicates Child Behavior Checklist; CI, confidence interval; FA, fractional anisotropy.

**Supplementary Table 1**. *MFD Sensitivity Analysis: APE Effects on White Matter FA With and Without Mean Framewise Displacement Covariate*

| **Tract** | **Primary model** | | **+MFD covariate** | | **Δ Estimate (%)** | **Sig. change?** |
|---|---|---|---|---|---|---|
| | ***b*** | ***p*** | ***b*** | ***p*** | | |
| *Uncinate fasciculus (LH)* | −0.00096 | .001 | −0.00087 | .001 | 8.8% | No |
| *Corpus callosum* | −0.00078 | .001 | −0.00069 | .001 | 11.3% | No |
| *Uncinate fasciculus (RH)* | −0.00071 | .005 | −0.00062 | .006 | 12.2% | No |
| *CgC (LH)* | −0.00087 | .031 | −0.00075 | .044 | 13.4% | No |
| *pSLF (RH)* | −0.00052 | .031 | −0.00044 | .041 | 15.7% | No |
| *SLF (RH)* | −0.00047 | .047 | −0.00040 | .064 | 16.6% | YES |
| *pSLF (LH)* | −0.00046 | .065 | −0.00037 | .093 | 18.9% | No |
| *CgC (RH)* | −0.00065 | .095 | −0.00055 | .135 | 15.9% | No |
| *tSLF (RH)* | −0.00027 | .250 | −0.00020 | .356 | 26.5% | No |
| *SLF (LH)* | −0.00025 | .301 | −0.00017 | .431 | 31.3% | No |
| *tSLF (LH)* | −0.00015 | .547 | −0.00007 | .749 | 51.6% | No |

*Note.* Primary model: APE + standard covariates. Sensitivity model: APE + standard covariates + mean framewise displacement (MFD; excluding y-displacement). Δ Estimate = percentage change in the APE coefficient after adding MFD. Sig. change? = whether significance status ($p < .05$) changed between models. Tracts ordered by primary model p value.

**Supplementary Table 2**. *Image-Quality (Frame-Censoring) Sensitivity Analyses for Baseline and Longitudinal APE Associations*

**A. Baseline FA associations, with and without adjustment for frame censoring**

| Tract | b (primary) | P (primary) | b (+ censoring) | P (+ censoring) |
|---|---|---|---|---|
| Uncinate fasciculus (LH) | −0.00096 | .001 | −0.00085 | .001 |
| Corpus callosum | −0.00079 | .001 | −0.00069 | .002 |
| Uncinate fasciculus (RH) | −0.00071 | .005 | −0.00061 | .011 |

**B. APE effect on FA across visits (censoring adjusted), with and without the APE × age interaction**

| Tract | b (no interaction) | P (no interaction) | b (with interaction) | P (with interaction) |
|---|---|---|---|---|
| Corpus callosum | −0.0149 | .004 | −0.0143 | .006 |
| Uncinate fasciculus (LH) | −0.0136 | .011 | −0.0130 | .016 |
| Cingulate (LH) | −0.0075 | .198 | −0.0061 | .303 |
| PSLF (RH) | −0.0069 | .213 | −0.0056 | .317 |
| SLF (RH) | −0.0069 | .221 | −0.0057 | .315 |
| Uncinate fasciculus (RH) | −0.0060 | .245 | −0.0050 | .331 |
| PSLF (LH) | −0.0057 | .301 | −0.0049 | .379 |
| Cingulate (RH) | −0.0053 | .390 | −0.0046 | .451 |
| TSLF (RH) | −0.0037 | .526 | −0.0031 | .606 |
| SLF (LH) | −0.0020 | .725 | −0.0012 | .829 |
| TSLF (LH) | 0.0002 | .968 | 0.0009 | .882 |

*Note.* APE burden was not associated with the number of censored frames at baseline ($b = 0.03$; $P = .31$); mean censored frames were 4.19, 4.28, 4.38, and 4.53 for burden levels 0, 1, 2, and 3+, respectively. In Part B, FA was z-scored within tract. All models adjusted for caregiver educational attainment, sex, race, ethnicity, age, MRI scanner software version, and (for longitudinal models) time-varying frame censoring, with random effects for participants nested within families and sites. b indicates unstandardized coefficient; FA, fractional anisotropy; APE, adverse prenatal exposure; LH, left hemisphere; RH, right hemisphere; PSLF, parietal superior longitudinal fasciculus; SLF, superior longitudinal fasciculus; TSLF, temporal superior longitudinal fasciculus.

**Supplementary Table 3**. *Maternal Depression Sensitivity Analysis: APE Effects on White Matter FA With and Without Adjustment for Biological-Mother Depression History*

**A. Baseline FA associations (full sample, N = 10,483; missingness modeled as a category)**

| Tract | b (primary) | P_FDR (primary) | b (+ mat. dep.) | P_FDR (+ MD) | Δ (%) |
|---|---|---|---|---|---|
| *Uncinate fasciculus (LH)* | −0.00096 | .011 | −0.00086 | .021 | 3.3 |
| *Corpus callosum* | −0.00079 | .011 | −0.00069 | .021 | 5.4 |
| *Uncinate fasciculus (RH)* | −0.00071 | .039 | −0.00062 | .054 | 3.5 |

**B. Baseline FA associations (complete-case, N = 5,387; known maternal-depression status only)**

| Tract | b | P_FDR |
|---|---|---|
| *Uncinate fasciculus (LH)* | −0.00130 | .003 |
| *Corpus callosum* | −0.00118 | .003 |
| *Uncinate fasciculus (RH)* | −0.00127 | .003 |

**C. APE × Age interaction (trajectory), with and without adjustment**

| Tract (APE × Age) | b (primary) | b (+ MD) | Δ (%) | Sig. change? |
|---|---|---|---|---|
| *Uncinate fasciculus (RH)* | 0.0040 | 0.0040 | 0.10 | No |
| *Cingulate cingulum (LH)* | 0.0040 | 0.0040 | −0.01 | No |
| *pSLF (RH)* | 0.0030 | 0.0030 | 0.14 | No |
| *Uncinate fasciculus (LH)* | 0.0028 | 0.0028 | −0.01 | No |
| *SLF (RH)* | 0.0026 | 0.0026 | 0.13 | No |

*Note.* Maternal depression history was drawn from the baseline family-history assessment (biological mother). Full-sample analyses (Part A) included all 10,483 participants, with missing maternal-depression status (n = 5,096) modeled as a separate category. Complete-case analyses (Part B) were restricted to the 5,387 participants with known status (no, n = 3,011; yes, n = 2,376). Because this item was unavailable for approximately half the sample, missingness was modeled as a separate category in the full-sample analysis (Part A) and confirmed in a complete-case analysis restricted to participants with known status (Part B); effects were comparable and, in the complete-case subsample, modestly stronger. Maternal depression history was positively associated with APE burden. Across all analyses, associations were attenuated by less than 6%

and trajectory interactions shifted by less than 0.3% with no change in significance status, indicating that associations between prenatal APE burden and FA were not attributable to maternal depression. Δ = percentage change in the APE coefficient after adding maternal depression (MD). APE indicates adverse prenatal exposure; FA, fractional anisotropy; FDR, false discovery rate; LH, left hemisphere; RH, right hemisphere; pSLF, parietal superior longitudinal fasciculus; SLF, superior longitudinal fasciculus.

**Supplementary Table 4**. *PRS Total Problems and Prenatal APE Main Effects on White Matter FA (European-Ancestry Subsample)*

| Tract | PRS b | PRS 95% CI; P | APE b | APE 95% CI; P |
|---|---|---|---|---|
| *CgC (LH)* | −0.0012 | −0.0023 to −0.0001; .030 | −0.00003 | −0.0013 to 0.0012; .967 |
| *pSLF (LH)* | −0.0006 | −0.0013 to 0.0000; .062 | −0.0006 | −0.0014 to 0.0001; .106 |
| *Corpus callosum* | −0.0006 | −0.0012 to 0.0001; .074 | −0.0006 | −0.0013 to 0.0001; .109 |
| *pSLF (RH)* | −0.0005 | −0.0012 to 0.0001; .118 | −0.0007 | −0.0014 to 0.00004; .062 |
| *CgC (RH)* | −0.0007 | −0.0018 to 0.0004; .214 | −0.0004 | −0.0016 to 0.0008; .512 |
| *SLF (RH)* | −0.0004 | −0.0011 to 0.0002; .219 | −0.0006 | −0.0013 to 0.0001; .096 |
| *SLF (LH)* | −0.0004 | −0.0011 to 0.0003; .235 | −0.0004 | −0.0011 to 0.0003; .268 |
| *Uncinate fasciculus (LH)* | −0.0004 | −0.0012 to 0.0004; .312 | −0.0006 | −0.0014 to 0.0003; .183 |
| *tSLF (LH)* | −0.0002 | −0.0009 to 0.0004; .492 | −0.0003 | −0.0011 to 0.0004; .377 |
| *Uncinate fasciculus (RH)* | −0.0001 | −0.0008 to 0.0006; .771 | −0.0002 | −0.0010 to 0.0005; .539 |
| *tSLF (RH)* | < .001 | −0.0007 to 0.0006; .926 | −0.0003 | −0.0011 to 0.0004; .369 |

*Note.* Linear mixed effects models in participants of European ancestry with available polygenic risk score data (n ≈ 6,270). Both PRS total problems (z-scored) and prenatal APE were included simultaneously as fixed effects, along with age, sex, race, ethnicity, caregiver educational attainment, MRI scanner model, and 10 ancestry principal components. Random effects: (1 | site / family_id). No PRS or APE effect survived FDR correction across the 11 tracts; the nominal PRS association with left cingulate FA (P = .030) did not survive correction. APE effects are non-significant in this subsample consistent with reduced power relative to the full analytic sample. Tracts ordered by PRS p value.

**Supplementary Table 5.** *Exploratory Domain-Specific Associations Between Individual Prenatal Exposures and White Matter Fractional Anisotropy*

**Part A. Baseline (cross-sectional) associations**

| Prenatal exposure | Tract | *b* | 95% CI | *P* |
|---|---|---|---|---|
| Alcohol | Corpus callosum | .00050 | −.00076, .00176 | .440 |
| Alcohol | Uncinate, left | −.00031 | −.00179, .00118 | .686 |
| Alcohol | Uncinate, right | .00012 | −.00123, .00147 | .860 |
| Birth complication | Corpus callosum | −.00106 | −.00228, .00017 | .091 |
| Birth complication | Uncinate, left | −.00181 | −.00324, −.00038 | .013 |
| Birth complication | Uncinate, right | −.00114 | −.00245, .00017 | .088 |
| Cannabis | Corpus callosum | −.00312 | −.00543, −.00081 | .008 |
| Cannabis | Uncinate, left | −.00268 | −.00537, .00001 | .051 |
| Cannabis | Uncinate, right | −.00196 | −.00442, .00050 | .118 |
| Pregnancy complication | Corpus callosum | −.00191 | −.00301, −.00082 | <.001 |
| Pregnancy complication | Uncinate, left | −.00171 | −.00299, −.00043 | .009 |
| Pregnancy complication | Uncinate, right | −.00161 | −.00278, −.00045 | .007 |
| Tobacco/nicotine | Corpus callosum | −.00073 | −.00237, .00091 | .382 |
| Tobacco/nicotine | Uncinate, left | −.00200 | −.00391, −.00008 | .041 |
| Tobacco/nicotine | Uncinate, right | −.00060 | −.00235, .00115 | .502 |
| Unplanned pregnancy | Corpus callosum | −.00077 | −.00193, .00039 | .194 |
| Unplanned pregnancy | Uncinate, left | −.00038 | −.00173, .00097 | .580 |
| Unplanned pregnancy | Uncinate, right | −.00070 | −.00193, .00053 | .265 |

**Part B. Developmental trajectory (exposure × age interaction) associations**

| Prenatal exposure | Tract | *b* | 95% CI | *P* |
|---|---|---|---|---|
| Alcohol | Cingulate cingulum, left | .00879 | .00179, .01580 | .014 |
| Alcohol | pSLF, right | .00518 | −.00092, .01130 | .096 |
| Alcohol | SLF, right | .00448 | −.00155, .01050 | .146 |
| Alcohol | Uncinate, left | .00096 | −.00540, .00732 | .767 |
| Alcohol | Uncinate, right | .01120 | .00523, .01720 | <.001 |
| Birth complication | Cingulate cingulum, left | .00244 | −.00447, .00935 | .488 |
| Birth complication | pSLF, right | −.00154 | −.00756, .00449 | .617 |
| Birth complication | SLF, right | −.00152 | −.00749, .00444 | .617 |
| Birth complication | Uncinate, left | .00038 | −.00592, .00668 | .906 |
| Birth complication | Uncinate, right | .00187 | −.00410, .00785 | .538 |
| Cannabis | Cingulate cingulum, left | .01570 | .00250, .02900 | .020 |

| | | | | |
|---|---|---|---|---|
| Cannabis | pSLF, right | .01180 | .00025, .02340 | .045 |
| Cannabis | SLF, right | .01040 | −.00108, .02190 | .076 |
| Cannabis | Uncinate, left | .01420 | .00213, .02630 | .021 |
| Cannabis | Uncinate, right | .01550 | .00408, .02690 | .008 |
| Pregnancy complication | Cingulate cingulum, left | .00207 | −.00409, .00823 | .511 |
| Pregnancy complication | pSLF, right | .00503 | −.00035, .01040 | .067 |
| Pregnancy complication | SLF, right | .00464 | −.00069, .00996 | .088 |
| Pregnancy complication | Uncinate, left | .00324 | −.00238, .00887 | .258 |
| Pregnancy complication | Uncinate, right | .00296 | −.00238, .00829 | .277 |
| Tobacco/nicotine | Cingulate cingulum, left | −.00158 | −.01070, .00755 | .735 |
| Tobacco/nicotine | pSLF, right | .00305 | −.00491, .01100 | .453 |
| Tobacco/nicotine | SLF, right | .00288 | −.00501, .01080 | .474 |
| Tobacco/nicotine | Uncinate, left | .00186 | −.00644, .01020 | .660 |
| Tobacco/nicotine | Uncinate, right | .00516 | −.00270, .01300 | .198 |
| Unplanned pregnancy | Cingulate cingulum, left | .01050 | .00411, .01690 | .001 |
| Unplanned pregnancy | pSLF, right | .00716 | .00159, .01270 | .012 |
| Unplanned pregnancy | SLF, right | .00655 | .00103, .01210 | .020 |
| Unplanned pregnancy | Uncinate, left | .00439 | −.00143, .01020 | .140 |
| Unplanned pregnancy | Uncinate, right | .00337 | −.00215, .00889 | .231 |

*Note.* All tested associations are reported, including nonsignificant effects. Part A reports cross-sectional (baseline) associations between each prenatal exposure and tract-averaged FA; Part B reports exposure × age interactions from longitudinal mixed-effects models. Each model included a single prenatal exposure and the full covariate set (child age, sex, race, ethnicity, caregiver educational attainment, and MRI scanner software version), with random effects for participant, family, and site. These analyses are exploratory and uncorrected for multiple comparisons. b indicates unstandardized regression coefficient; CI, confidence interval; FA, fractional anisotropy; pSLF, parietal superior longitudinal fasciculus; SLF, superior longitudinal fasciculus.

**Supplementary Table 6**. *Mediation Analyses: Prenatal APE → White Matter FA → Emotional Dysregulation Across 11 A Priori Tracts*

| Tract | Path a | | | Path b | | | Indirect (a×b) | 95% CI | |
|---|---|---|---|---|---|---|---|---|---|
| | ***b*** | **SE** | ***p*** | ***b*** | **SE** | ***p*** | | | |
| *Uncinate fasciculus (LH)* | −0.023 | 0.006 | .000 | −0.057 | 0.012 | < .001 | 0.00129 | [0.00026, 0.00289] | * |
| *Uncinate fasciculus (RH)* | −0.017 | 0.006 | .006 | −0.051 | 0.013 | < .001 | 0.00086 | [0.00009, 0.00195] | * |
| *Corpus callosum* | −0.019 | 0.006 | .002 | −0.053 | 0.013 | < .001 | 0.00098 | [0.00015, 0.00278] | * |
| *CgC (LH)* | −0.017 | 0.007 | .012 | −0.025 | 0.011 | .025 | 0.00044 | [−0.00044, 0.00133] | |
| *pSLF (RH)* | −0.014 | 0.007 | .034 | −0.046 | 0.012 | < .001 | 0.00064 | [−0.00013, 0.00176] | |
| *SLF (RH)* | −0.013 | 0.007 | .052 | −0.043 | 0.012 | < .001 | 0.00055 | [−0.00020, 0.00165] | |
| *pSLF (LH)* | −0.012 | 0.007 | .067 | −0.049 | 0.012 | < .001 | 0.00059 | [−0.00013, 0.00166] | |
| *CgC (RH)* | −0.010 | 0.007 | .172 | −0.028 | 0.011 | .010 | 0.00028 | [−0.00020, 0.00105] | |
| *SLF (LH)* | −0.008 | 0.007 | .213 | −0.045 | 0.012 | < .001 | 0.00037 | [−0.00024, 0.00136] | |
| *tSLF (RH)* | −0.007 | 0.007 | .264 | −0.035 | 0.012 | .004 | 0.00026 | [−0.00043, 0.00107] | |
| *tSLF (LH)* | −0.006 | 0.007 | .379 | −0.042 | 0.012 | < .001 | 0.00025 | [−0.00034, 0.00118] | |

*Note.* Bootstrap mediation analyses (n = 1,000 iterations). Path a = APE → FA; path b = FA → dysregulation (controlling for APE). Indirect effect = path a × path b. Total effect of APE on dysregulation: b = 0.131, SE = 0.008, $p < 10^{-54}$. Direct effect of APE on dysregulation (path c') remained significant across all tracts after accounting for FA (all $p < 10^{-49}$), indicating partial mediation where significant. FA values z-scored. 95% CI = percentile bootstrap confidence interval. * = CI excludes zero (significant indirect effect). CgC = cingulum cingulate; LH = left hemisphere; RH = right hemisphere; pSLF = parietal superior longitudinal fasciculus; SLF = superior longitudinal fasciculus; tSLF = temporal superior longitudinal fasciculus.
